# AI Decision Support for Challenging Teledermatology Cases: MedGemma Performance in the Dermatology ECHO Program

**DOI:** 10.64898/2026.05.21.26353523

**Authors:** Jeffrey B. Appiagyei, Ruth O. Otu, Mollie Henry, Benjamin W. Casterline, Mirna Becevic

## Abstract

Teledermatology expands access to dermatologic expertise in rural settings, yet diagnostic uncertainty persists in low-resource primary care. This retrospective study evaluated MedGemma-4B-IT, a compact multimodal vision-language model, as adjunctive clinical decision support for challenging diagnostic cases. We analyzed 77 zero-concordance cases (360 clinical photographs) from a Dermatology Extension for Community Healthcare Outcomes (ECHO) tele-mentoring program (2016-2021). Zero-concordance cases showed no overlap between primary clinician provisional diagnosis and dermatologist-confirmed diagnosis. The model was prompted using dermatologist-style format to generate ranked differential diagnoses. Performance was assessed using strict case-level top-k exact-match accuracy and relaxed matching criteria based on fuzzy string similarity. MedGemma achieved 0.0% strict top-1 accuracy, 1.3% top-3 accuracy, 3.9% top-5 accuracy, and 3.9% top-10 accuracy. Relaxed concept-level matching achieved 28.6% top-1, 63.6% top-5, and 67.5% top-10 accuracy. Image-level accuracy was 44.2% (159/360, 95% CI 39.0-49.5%). The model surfaced the correct diagnosis within differential lists in 45.5% of cases despite no exact top-1 matches, suggesting utility for differential expansion rather than definitive diagnosis. Performance varied across diagnostic categories, with highest accuracy in Other categories (54.5%) and lowest in neoplastic conditions (0.0%). Common errors included confusion between inflammatory and other diagnostic groupings. These findings characterize MedGemma performance on real-world teledermatology cases and inform safe, clinician-in-the-loop integration into teledermatology workflows where specialist oversight remains essential.

**What this Study Adds:** This study provides empirical evaluation of MedGemma-4B-IT as adjunctive decision support for challenging teledermatology cases in a community healthcare ECHO setting. We demonstrate that while strict top-1 diagnostic accuracy is 0%, the model correctly surfaces the dermatologist-confirmed diagnosis within a 10-item differential in 45.5% of zero-concordance cases, suggesting value as a differential diagnostic prompt rather than a direct diagnostic replacement. These findings inform safe, clinician-in-the-loop deployment strategies for compact vision-language models in resource-limited telemedicine settings.

**Conclusions:** MedGemma demonstrates differential diagnostic utility in challenging teledermatology cases, surfacing the correct diagnosis within a 10-item differential in nearly half of cases despite zero top-1 accuracy. These findings support clinician-in-the-loop AI deployment for diagnostic expansion in resource-limited settings, while highlighting the need for improved neoplastic detection and confidence calibration in future model development.

**Results:** Under strict exact matching, top-1 accuracy was 0.0% (0/77), increasing to 3.9% (3/77) at top-10. Under relaxed concept-level matching, top-1 accuracy was 28.6% (22/77), rising to 45.5% (35/77) at top-10. The Mean Reciprocal Rank was 0.4287. Diagnostic performance varied by category: Other diagnoses showed 54.5% top-10 accuracy, while neoplastic conditions showed 0.0%.

**Methods:** This retrospective study analyzed 77 zero-concordance cases (360 images) from the Missouri Dermatology ECHO program (2016-2021). Cases were those where the primary care clinician provisional diagnosis showed no textual overlap with the dermatologist-confirmed diagnosis. Primary outcome was top-1 exact-match accuracy; secondary outcomes included top-k accuracy under concept-level relaxed matching.

**Background:** Teledermatology expands access to dermatologic expertise in rural settings, yet diagnostic uncertainty persists in low-resource primary care. We evaluated MedGemma-4B-IT, a compact multimodal vision-language model, as adjunctive clinical decision support for challenging teledermatology cases.

- Stratified performance analysis by diagnostic category and image count provides actionable guidance for deployment scenarios.
- Concept-level relaxed matching reveals clinically relevant differential diagnostic utility that strict exact-match metrics obscure.
- Zero-concordance cases provide a rigorous test of model performance, representing the diagnostic frontier where clinical decision support is most needed.

## 1. Introduction

Teledermatology has matured into an evidence-based pathway for expanding access to dermatologic expertise, particularly for rural and underserved populations [1,2]. Systematic reviews demonstrate clinically useful diagnostic and management outcomes across diverse settings [1,3]. However, teledermatology introduces sources of variability: heterogeneous image quality, incomplete clinical documentation, missing physical examination context, and diagnostic uncertainty among primary care providers [4]. Diagnostic discordance between non-dermatologists and dermatologists is well documented [4]. Clinicians and telehealth programs benefit from transparent, rigorously evaluated decision support tools that integrate seamlessly into clinical workflows [5,6].

**Figure 1:**
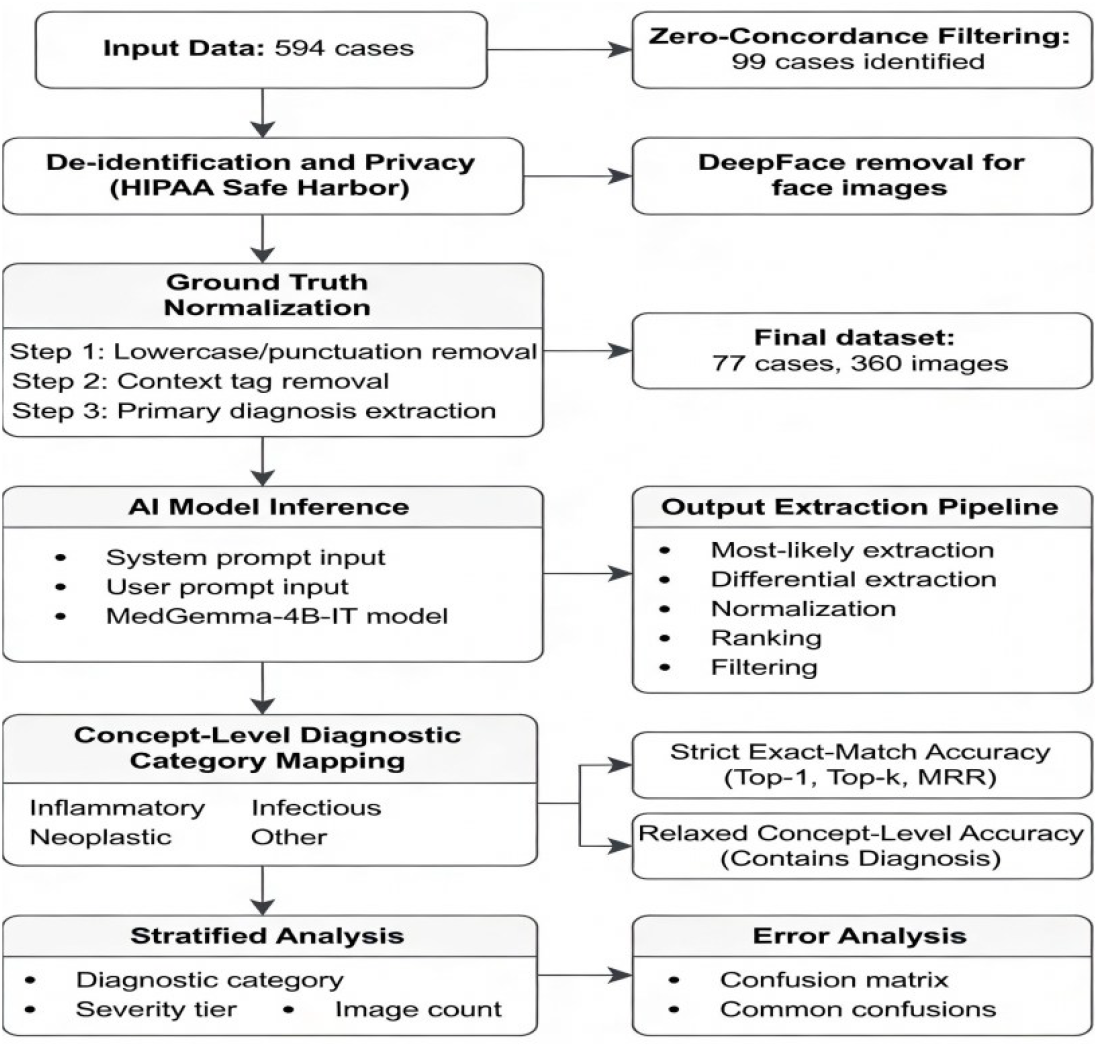
Study workflow.. The figure illustrates the complete analytical process from initial cohort selection (594 cases) through identification of zero-concordance cases (99 cases), de-identification using DeepFace, diagnostic normalization, MedGemma inference, and accuracy evaluation.

Recent advances in foundation models have created a new class of multimodal decision support tools capable of jointly reasoning over clinical text and images [7,8]. However, they are prone to hallucination and must be carefully evaluated for reliability and safety in clinical contexts before deployment [9,10]. In dermatology, prior image-based artificial intelligence (AI) systems have achieved dermatologist-level performance for specific classification tasks under controlled laboratory conditions [9]. Yet real-world teledermatology cases often present heterogeneous lesions, variable skin tones, and incomplete metadata, conditions that can substantially degrade performance and raise important equity concerns [11].

Zero-concordance cases, in which the primary clinician provisional diagnosis shows no overlap with the dermatologist-confirmed diagnosis, represent the diagnostic frontier where decision support is most needed [12]. These cases often involve atypical presentations of common conditions, rare diagnoses unfamiliar to primary care providers, or conditions whose visual appearance varies substantially across skin tones. For patients in rural or underserved areas where dermatologist access is limited, diagnostic delays in these challenging cases can lead to inappropriate treatment, unnecessary biopsies, or progression of time-sensitive conditions.

This study evaluates MedGemma-4B-IT, a compact multimodal vision language model, as adjunctive clinical decision support for zero-concordance teledermatology presentations [13]. We quantify diagnostic performance using strict and relaxed matching criteria, characterize common error patterns, and provide guidance for safe, clinician-in-the-loop deployment.

## 2. Methods

### A. Study Design and Setting

This was a retrospective, observational evaluation of de-identified dermatology Extension for Community Healthcare Outcomes (ECHO) cases submitted by community-based clinicians through the Missouri Telehealth Network (Show-Me ECHO) tele-mentoring project. The analysis focuses on zero-concordance cases, defined as instances in which the submitting clinician diagnosis and the dermatologist final diagnosis exhibited no overlap after text normalization.

### B. Case Selection and Cohort Construction

This study drew from 594 tele-ECHO cases submitted between 2016 and 2021. Stringent criteria identified zero-concordance cases through text normalization including lowercasing and punctuation removal [12]. This filtering identified 99 cases. Automated face detection was applied, followed by removal using DeepFace to exclude cases containing potentially identifiable facial features. The final analytic dataset comprised 77 cases and 360 clinical images. Each case consisted of a structured clinical vignette (patient age, skin type when documented, lesion location, symptoms, symptom duration, relevant history) paired with one or more clinical photographs.

### C. De-identification and Privacy Protection

All case materials underwent de-identification following HIPAA Safe Harbor principles. Direct identifiers are not collected in tele-ECHO. Free-text clinical vignettes underwent automated string matching and manual review. Facial photographs were excluded using automated face-detection pipeline (DeepFace) with manual verification.

### D. Ground Truth Labeling and Diagnostic Normalization

The dermatologist-confirmed final diagnosis served as the authoritative ground truth label. Free-text diagnoses were normalized through systematic three-step process: (1) lowercasing and removal of punctuation and extra whitespace; (2) rule-based removal of clinical context tags; and (3) extraction of the primary diagnosis term (first diagnosis before commas or slashes). Normalized diagnoses were mapped post hoc to broader clinical diagnostic groups (Inflammatory, Infectious, Neoplastic, Other) for stratified analysis.

### E. Model Selection and Prompting Strategy

This study evaluated MedGemma-4B-IT, a compact multimodal vision language model trained on medical data and optimized for instruction following [13]. A dermatologist-style prompting approach was used to elicit ranked diagnostic candidates.

#### System Prompt

“You are a board certified dermatologist. You will be shown multiple images of a skin condition for a single patient.”

#### User Prompt

“Based on all the images provided and the clinical context, provide the most likely diagnosis and up to two differential diagnoses ranked by likelihood.”

### F. Inference and Output Extraction

For each case, all clinical images and the normalized vignette were passed to MedGemma-4B-IT. The model generated free-text outputs, which were post-processed using a multi-stage extraction pipeline: (1) Most-Likely Extraction; (2) Differential Extraction; (3) Normalization; (4) Ranking; and (5) Filtering of generic terms.

### G. Outcome Definitions and Accuracy Metrics

Primary endpoint was strict case-level top-1 exact-match accuracy. A case was counted as correct only when the normalized top-1 predicted diagnosis exactly matched the normalized ground truth diagnosis. Secondary endpoints included top-k exact-match accuracy for k = 3, 5, and 10. Mean reciprocal rank (MRR) was computed at the case level. Concept-level performance was evaluated by mapping diagnoses to broader diagnostic categories.

### H. Stratified Analyses

Performance was summarized across three stratification dimensions: diagnostic categories, clinical severity tiers (benign, other, serious), and number of clinical images per case (1-2, 3-4, 5+ images).

### I. Statistical Methods

This study reports proportions with 95% confidence intervals computed using the binomial Wilson score method. Mean Reciprocal Rank (MRR) was computed as: MRR = (1/N) x Sigma (1/rank) where rank is the position of the first correct diagnosis. Top-k accuracy was defined as: Accuracy(k) = [Number of cases where correct diagnosis is in Top-k predictions / N] x 100. All analyses were conducted in Python using reproducible scripts on the University of Missouri Hellbender HPC cluster [14].

## 3. Results

### A. Cohort Characteristics

The final analytic cohort consisted of 77 zero-concordance teledermatology cases with 360 clinical images. The number of images per case ranged from 1 to 17 (mean = 4.68, SD = 3.30).

Under strict exact matching, top-1 accuracy was 0.0%, increasing to 1.3% at top-3 and 3.9% at top-5 and top-10. Concept-level relaxed matching achieved 28.6% at top-1 and increased substantially with larger k, reaching 63.6% at top-5 and 67.5% at top-10. Mean reciprocal rank (concept, k<=10) was 0.43. Image-level accuracy was 44.2% (159/360, 95% CI 39.0-49.5%). Human-reviewed Contains Diagnosis criterion showed 45.5% (35/77) of cases contained the correct diagnosis within model output.

The striking discrepancy between strict top-1 accuracy (0.0%) and top-10 accuracy (45.5%) highlights a key finding: MedGemma rarely identifies the correct diagnosis as its first choice but frequently includes it within an expanded differential list. This pattern suggests the model utility may lie primarily in differential expansion and triage support rather than in replacement of clinician judgment.

**Table 1:** Cohort Characteristics and Diagnostic Accuracy Metrics.

| Metric | Value (n, %) |
| --- | --- |
| Cases (N) | 77 |
| Images (N) | 360 |
| Mean images per case (SD) | 4.68 (3.30) |
| Strict Top-1 exact match | 0/77 (0.0%) |
| Strict Top-10 exact match | 3/77 (3.9%) |
| Concept Top-1 match | 22/77 (28.6%) |
| Concept Top-10 match | 52/77 (67.5%) |
| Mean Reciprocal Rank (concept, $k \leq 10$) | 0.4287 |
| Human-reviewed Contains Diagnosis | 35/77 (45.5%) |

### B. Accuracy by Diagnostic Category

Diagnostic performance varied substantially across clinical categories. The Other category showed the highest top-10 accuracy (54.5%, 18/33). Inflammatory conditions achieved 21.7% (5/23) accuracy. Autoimmune conditions showed 66.7% (2/3) accuracy. Benign conditions achieved 44.4% (4/9) accuracy. The complete failure on neoplastic conditions (0.0%, 0/2) is clinically concerning.

**Table 2:** Top-10 Accuracy by Diagnostic Category.

| Diagnostic Category | N | Top-10 Accuracy | 95% CI |
| --- | --- | --- | --- |
| Inflammatory | 23 | 21.7% (5/23) | 7.5-43.7% |
| Infectious | 3 | 33.3% (1/3) | 0.8-90.6% |
| Autoimmune | 3 | 66.7% (2/3) | 9.4-99.2% |
| Benign | 9 | 44.4% (4/9) | 13.7-78.8% |
| Other | 33 | 54.5% (18/33) | 36.4-71.9% |
| Neoplastic | 2 | 0.0% (0/2) | 0.0-84.2% |

### C. Performance by Image Count

Cases with 3-4 images showed the highest accuracy (50.0%, 11/22), while cases with 1-2 images showed 44.4% (4/9) and cases with 5+ images showed 43.5% (20/46). The Pearson correlation between image count and case-level correctness was near zero (r = -0.029, p = 0.803).

**Table 3:** Performance by Image Count Group.

| Image Count Group | N Cases | Top-10 Accuracy | 95% CI | Mean MRR |
| --- | --- | --- | --- | --- |
| 1-2 images | 9 | 44.4% | 13.7-78.8% | 0.5714 |
| 3-4 images | 22 | 50.0% | 28.2-71.8% | 0.4167 |
| 5+ images | 46 | 43.5% | 29.0-58.9% | 0.3392 |

### D. Category-Level Confusion Analysis

The dominant confusion pattern involved misclassification of Other diagnoses into the Infectious category (n = 10). Inflammatory conditions such as dermatitis or eczema were frequently misclassified as Other (n = 7). These systematic category-level errors highlight specific targets for future model refinement.

### E. Diagnostic Match Summary

Using fuzzy string matching with manual adjudication, there were no strict exact matches (0.0%) and no highly similar matches (0.0%). Under relaxed matching, the correct diagnosis appeared somewhere in the model output for 35 of 77 cases (45.5%), while 42 of 77 cases (54.5%) showed no match.

**Table 4:** Common Category-Level Diagnostic Confusions.

| Ground Truth | Predicted Category | N |
| --- | --- | --- |
| Other | Infectious | 10 |
| Dermatitis or eczema | Other | 7 |
| Other | Dermatitis or eczema | 4 |
| Other | Neoplastic | 4 |
| Lichen | Other | 2 |
| Other | Acne | 2 |

## 4. Discussion

### A. Key Findings and Clinical Interpretation

**Figure 2:**
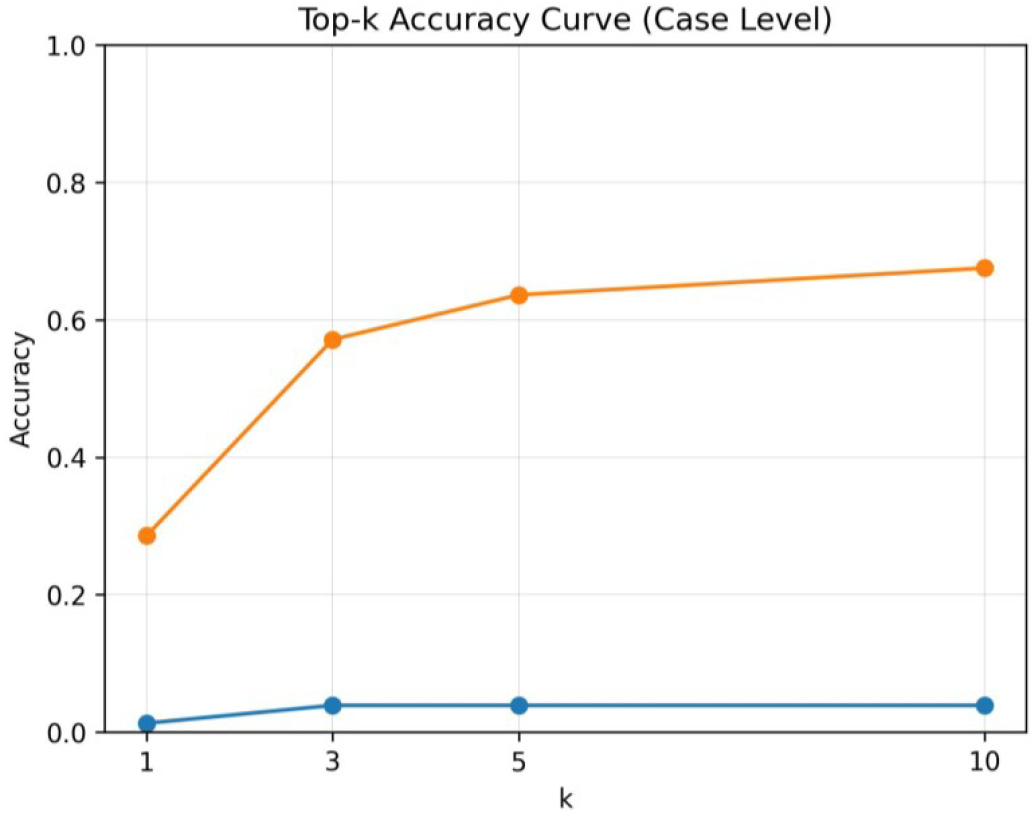
Top-k accuracy curves comparing strict exact-match vs. concept-level relaxed matching. The figure shows accuracy at increasing k values (1-10) for both matching criteria.

**Figure 3:**
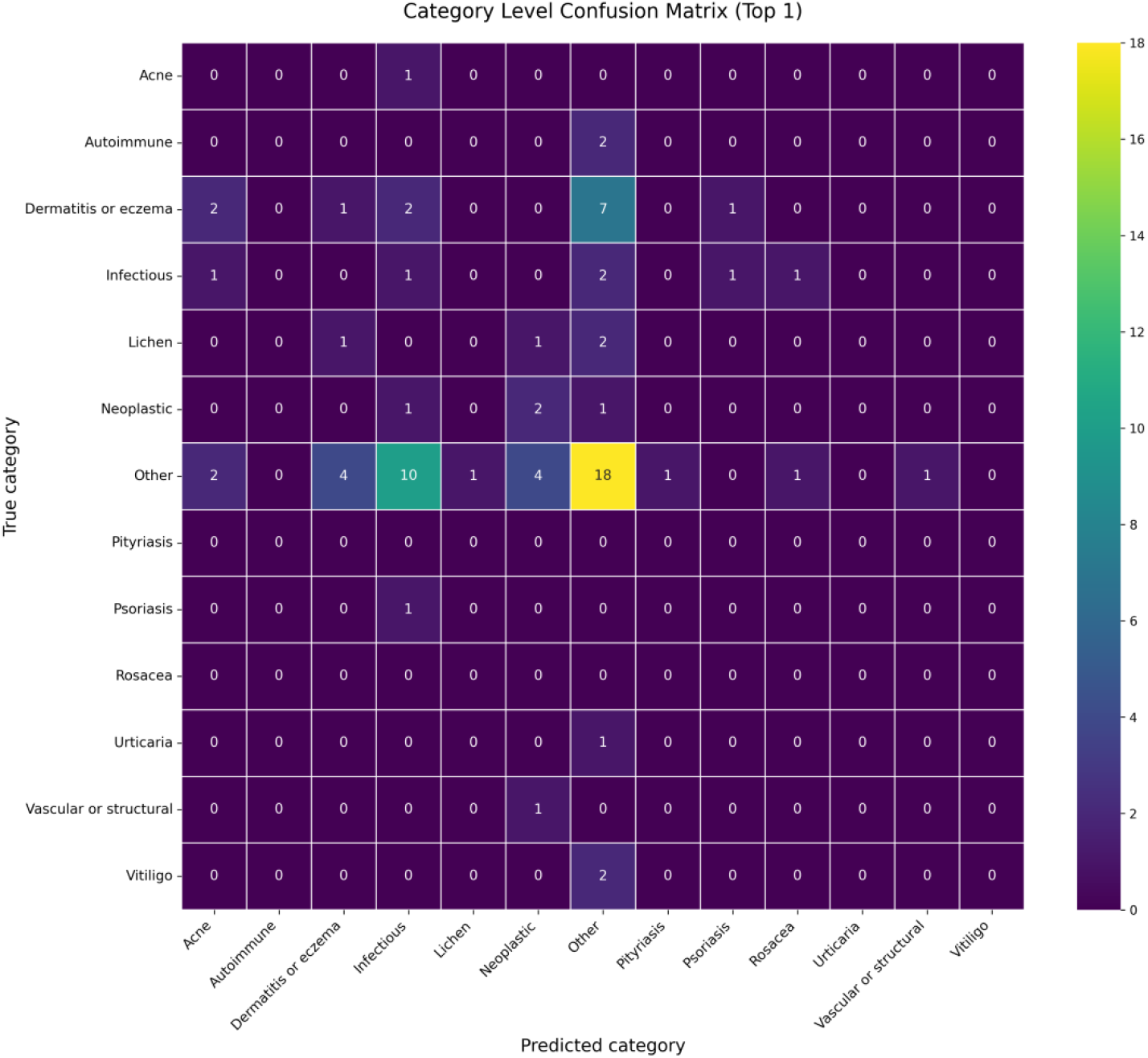
Diagnostic category confusion matrix. Rows represent ground truth categories; columns represent MedGemma-predicted categories. Color intensity indicates frequency of diagnostic misclassification between categories.

This retrospective evaluation reveals a nuanced performance profile that informs realistic expectations for multimodal AI in clinical practice. First, the model achieved 0.0% strict top-1 accuracy, yet the correct diagnosis appeared in 45.5% of cases under relaxed matching, suggesting the model functions more reliably as a differential expansion aid. Second, performance varied across diagnostic categories. Third, image count did not predict accuracy (r = -0.029). Fourth, the most frequent error was misclassification of Other diagnoses as Infectious (n = 10).

The 45.5% rate at which MedGemma surfaced the correct diagnosis suggests potential value as a second opinion prompt. However, the complete failure on neoplastic conditions (0/2 cases) raises critical safety concerns. The model tendency to over-predict infectious etiologies could lead to unnecessary antimicrobial prescribing.

### B. Context and Comparison with Prior Work

These results should be interpreted in the broader context of AI in dermatology and teledermatology. Prior dermatology AI studies often report strong performance in controlled settings, while real-world teledermatology commonly involves heterogeneous image quality [1,2,9,11]. The 45.5% relaxed Contains Diagnosis rate supports potential value as a second opinion signal rather than as a standalone diagnostic output.

### C. Safety and Implementation Considerations

For teledermatology, we recommend a clinician-in-the-loop model in which MedGemma outputs are presented as supporting information rather than as recommendations [15,16]. Clinicians should retain full diagnostic authority. Future deployments should incorporate calibrated confidence scores. This dataset included cases from diverse populations, yet the study did not conduct stratified performance analysis by race or skin tone.

### D. Implications for Teledermatology Workflows

MedGemma appears most suitable for cases of diagnostic ambiguity or low clinician confidence in primary care settings. Example use cases include: (1) rare or atypical presentations; (2) cases before specialist referral to triage urgency; (3) as a quality-improvement tool. The model is not recommended as a primary diagnostic tool.

Future Directions. Based on these findings, we propose several high-priority future investigations. First, fine-tuning MedGemma on a larger teledermatology cohort with balanced representation across diagnostic categories could improve neoplastic detection, which showed critical failures in this evaluation. Second, prospective studies should evaluate MedGemma in live clinical workflows to assess real-world usability, clinician trust, and impact on diagnostic decision-making. Third, comparative studies against other vision-language models and against unaided clinician performance would contextualize these results. Finally, deployment frameworks for safe, clinician-in-the-loop AI assistance in resource-limited settings represent a critical area for implementation research.

### E. Limitations

This study has several limitations. First, cases were drawn from a single ECHO program, which may limit generalizability. Second, the model was evaluated in a zero-shot setting without fine-tuning. Third, the analysis did not include saliency methods. Fourth, the study did not include head-to-head comparisons of other models.

## Supporting information

code_images

## Data Availability

All data produced in the present study are available upon reasonable request to the authors

## Statements and Declarations

Funding. This research did not receive any specific grant funding.

## Ethical Approval

This retrospective study was conducted under institutional quality-improvement protocols and does not constitute human subjects research requiring IRB approval. All case data were de-identified according to HIPAA Safe Harbor principles prior to analysis. Model inference was conducted on de-identified data only. No identifying information was included in any analysis, publication, or supplementary material.

## Conflict of Interest The authors declare no conflicts of interest

## Acknowledgments

The authors thank the Missouri Telehealth Network (Show-Me ECHO) for facilitating teledermatology cases for this study and the University of Missouri Hellbender HPC cluster support team for computational resources.

## Data Availability Statement

The data that support the findings of this study are not publicly available due to patient privacy protections under HIPAA Safe Harbor de-identification standards. De-identified aggregate data may be available from the corresponding author upon reasonable request and subject to institutional approvals.

