## Supplementary material for "AI Decision Support for Challenging Teledermatology Cases: MedGemma Performance in the Dermatology ECHO Program": code_images: MedGemma_Evaluation_Notebook.ipynb

```
# %% [markdown]
# MedGemma Teledermatology Evaluation Notebook

This notebook computes case level Top 1 exact match accuracy, Top k accuracy, MRR, performance by image count, a category level confusion matrix, and common category confusions.

Notes:
- This notebook treats empty or missing predictions as incorrect.
- Exact match is strict (string normalized).
- Concept match groups diagnoses into broader clinical buckets to make evaluation more robust to wording differences.


# %% [code]
import pandas as pd
import numpy as np
import re, os, math
from collections import Counter
from sklearn.metrics import confusion_matrix, classification_report
import matplotlib.pyplot as plt

input_path = r"/mnt/data/better_PromptMedGemma_AllECHO_Diagnoses_WITH_STATS3.xlsx"
df = pd.read_excel(input_path, sheet_name=0)
df.shape, df.columns.tolist()


# %% [code]
# Helper functions
def clean_text(x):
    if pd.isna(x):
        return ""
    x=str(x)
    return x.replace("\u2014","-").replace("\u2013","-")

def normalize_diag(x):
    x=clean_text(x).strip()
    parts=[p.strip() for p in re.split(r'[\r\n]+', x) if p.strip()]
    if parts:
        x=parts[0]
    x=re.sub(r'\bbx\b.*$', '', x, flags=re.I).strip()
    x=re.sub(r'\(.*?\)', '', x).strip()
    x=re.sub(r'\[.*?\]', '', x).strip()
    x=re.sub(r'\bcategory\b', '', x, flags=re.I).strip()
    x=re.sub(r'[:;,.]+$', '', x).strip()
    x=re.sub(r'\s+', ' ', x).strip().casefold()
    return x

def gt_primary(diag):
    x=clean_text(diag)
    parts=[p.strip() for p in re.split(r'[\r\n]+', x) if p.strip()]
    if parts:
        x=parts[0]
    x=re.sub(r'\bbx\b.*$', '', x, flags=re.I).strip()
    x=re.sub(r'\(.*?\)', '', x).strip()
    x=re.sub(r'\bcategory\b', '', x, flags=re.I).strip()
    x=re.split(r'\s*(?:,|/|;|\band\b|\bor\b)\s*', x, maxsplit=1, flags=re.I)[0].strip()
    x=re.sub(r'[:;,.]+$', '', x).strip()
    x=re.sub(r'\s+',' ', x).casefold()
    return x

diagnosis_kw = re.compile(r'(dermatitis|eczema|psoriasis|lichen|pityriasis|tinea|candid|impetigo|cellulitis|folliculitis|acne|rosacea|urticaria|vascul|lupus|pemph|pemphigoid|pemphigus|carcinoma|melanoma|keratosis|nevus|angioma|hemangioma|cyst|lipoma|sarcoma|granuloma|scabies|molluscum|zoster|herpes|wart|alopecia|ichthyosis|parapsoriasis|erythema|toxic epidermal|stevens-johnson|drug eruption|contact|seborrheic|hidradenitis|pyoderma|mycosis|necrolytic|acanthosis|blister)', re.I)
bad_kw = re.compile(r'^(appearance|location|assessment|differential diagnoses?|differential diagnosis|diagnosis|impression|history|recommendations?|plan|findings?|clinical|images?|important note|note|clinical presentation|physical exam)$', re.I)
adjective_kw = re.compile(r'(raised|redness|inflammation|bumps?|patch|plaque|lesion|rash|skin|area|scalp|face|trunk|extremit)', re.I)
generic_phrases = re.compile(r'(clinical presentation|physical exam|patient history|clinical history|key findings|management|treatment|recommend)', re.I)
blacklist_prefix = ("other","further","exposure","additional","consider","information","history","recommend")

def filter_diag2(n):
    if not n:
        return False
    if bad_kw.match(n):
        return False
    if generic_phrases.search(n):
        return False
    for p in blacklist_prefix:
        if n==p or n.startswith(p+" "):
            return False
    if diagnosis_kw.search(n):
        return True
    if len(n.split())>=2 and not adjective_kw.search(n):
        return True
    return False

def extract_all_most_likely(t):
    t=clean_text(t)
    out=[]
    for m in re.finditer(r'most likely diagnosis\s*:?\s*(.+)', t, flags=re.I):
        line=m.group(1)
        m2=re.search(r'\*\*(.+?)\*\*', line)
        if m2:
            out.append(m2.group(1))
        else:
            out.append(re.split(r'[.\n;]', line)[0])
    return out

def extract_all_differentials(t):
    t=clean_text(t)
    items=[]
    for m in re.finditer(r'differential diagnoses?\s*:?\s*', t, flags=re.I):
        start=m.end()
        tail=t[start:]
        cut=re.split(r'\n\s*(?:most likely diagnosis|recommendations?|plan|important note|next steps|additional information|clinical history)\s*:?'
                     , tail, flags=re.I)[0]
        lines=cut.splitlines()[:120]
        for line in lines:
            line=line.strip()
            if not line:
                continue
            mnum=re.match(r'^\s*(\d+)[\).\:-]\s*(.+)$', line)
            if mnum:
                items.append(mnum.group(2).strip())
                continue
            mbul=re.match(r'^\s*[-•]\s*(.+)$', line)
            if mbul:
                items.append(mbul.group(1).strip())
                continue
            if '**' in line:
                for m3 in re.finditer(r'\*\*(.+?)\*\*', line):
                    items.append(m3.group(1).strip())
    return items

def build_ranked_candidates(t, maxn=50):
    raw=[]
    raw += extract_all_most_likely(t)
    raw += extract_all_differentials(t)
    raw += [m.group(1) for m in re.finditer(r'\*\*(.+?)\*\*', clean_text(t))]
    norm=[]
    for item in raw:
        item=clean_text(item)
        item=re.sub(r'\*','',item).strip()
        item=re.split(r'\s*-\s*', item)[0].strip()
        item=re.split(r'\s*:\s*', item)[0].strip()
        item=re.split(r'\s*\(\s*', item)[0].strip()
        if len(item)<3:
            continue
        n=normalize_diag(item)
        if not filter_diag2(n):
            continue
        norm.append(n)
    if not norm:
        return []
    counts=Counter(norm)
    first={}
    for i,n in enumerate(norm):
        if n not in first:
            first[n]=i
    ranked=sorted(counts.keys(), key=lambda n:(-counts[n], first[n]))
    return ranked[:maxn]

def concept_key(diag):
    d=diag or ""
    if re.search(r'(dermatitis|eczema)', d): return "Dermatitis or eczema"
    if re.search(r'psoriasis', d): return "Psoriasis"
    if re.search(r'(tinea|fungal|candid|mycos|impetigo|cellulitis|folliculitis|herpes|zoster|wart|molluscum|scabies|infect)', d):
        return "Infectious"
    if re.search(r'(carcinoma|melanoma|bcc|scc|neoplasm|tumou?r|sarcoma|lymphoma)', d): return "Neoplastic"
    if re.search(r'(pemph|bullous)', d): return "Blistering"
    if re.search(r'(lupus|vascul|autoimmune)', d): return "Autoimmune"
    if re.search(r'(urticaria|hives)', d): return "Urticaria"
    if re.search(r'(lichen)', d): return "Lichen"
    if re.search(r'(pityriasis)', d): return "Pityriasis"
    if re.search(r'(rosacea)', d): return "Rosacea"
    if re.search(r'(acne)', d): return "Acne"
    if re.search(r'(alopecia)', d): return "Alopecia"
    if re.search(r'(vitiligo)', d): return "Vitiligo"
    if re.search(r'(angioma|hemangioma|vascular|cyst|lipoma|fibroma|keloid)', d): return "Vascular or structural"
    if d.strip()=="":
        return "Unknown"
    return "Other"

def img_group(n):
    if pd.isna(n): return "Unknown"
    try:
        n=float(n)
    except:
        return "Unknown"
    if n<=2: return "1 to 2"
    if n<=4: return "3 to 4"
    return "5 or more"


# %% [code]
# Standardize columns (edit these if your sheet uses different names)
df2 = df.rename(columns={
    "ECHO ID":"echo_id",
    "Final Diagnosis":"final_diagnosis",
    "AI Diagnosis Combined (MedGemma)":"ai_output",
    "Number of Images":"n_images",
    "Diagnosis Match Comment":"manual_match_comment"
}).copy()

df2["echo_id"]=df2["echo_id"].astype(str)
df2["gt_primary"]=df2["final_diagnosis"].apply(gt_primary)
df2["pred_list"]=df2["ai_output"].apply(build_ranked_candidates)
df2["top1_pred"]=df2["pred_list"].apply(lambda x: x[0] if isinstance(x,list) and len(x)>0 else "")

for k in [1,3,5,10]:
    df2[f"top{k}_exact"]=df2.apply(lambda r: int(r["gt_primary"] in (r["pred_list"][:k] if isinstance(r["pred_list"],list) else [])), axis=1)

df2["gt_key"]=df2["gt_primary"].apply(concept_key)
df2["pred_keys"]=df2["pred_list"].apply(lambda lst: [concept_key(x) for x in lst] if isinstance(lst,list) else [])
for k in [1,3,5,10]:
    df2[f"top{k}_concept"]=df2.apply(lambda r: int(r["gt_key"] in (r["pred_keys"][:k] if isinstance(r["pred_keys"],list) else []) and r["gt_key"]!="Unknown"), axis=1)

def mrr_concept(row, kmax=10):
    g=row["gt_key"]
    if g=="Unknown":
        return 0.0
    for i,pk in enumerate(row["pred_keys"][:kmax], start=1):
        if pk==g:
            return 1.0/i
    return 0.0

df2["mrr_concept"]=df2.apply(mrr_concept, axis=1)

df2["manual_contains"]=df2["manual_match_comment"].astype(str).str.contains("Contains Diagnosis", case=False, na=False).astype(int)
df2["img_group"]=df2["n_images"].apply(img_group)

df2.head()


# %% [code]
# Overall metrics
k_values=[1,3,5,10]
summary = pd.DataFrame({
    "Metric":[
        "Top 1 exact match",
        "Top 5 exact match",
        "Top 10 exact match",
        "Top 1 concept match",
        "Top 5 concept match",
        "Top 10 concept match",
        "MRR (concept, up to Top 10)",
        "Manual Contains Diagnosis rate"
    ],
    "Value":[
        df2["top1_exact"].mean(),
        df2["top5_exact"].mean(),
        df2["top10_exact"].mean(),
        df2["top1_concept"].mean(),
        df2["top5_concept"].mean(),
        df2["top10_concept"].mean(),
        df2["mrr_concept"].mean(),
        df2["manual_contains"].mean()
    ]
})
summary["Value_percent_or_value"]=summary["Value"].apply(lambda x: round(float(x)*100,2) if x<=1 else round(float(x),4))
summary


# %% [code]
# Top-k accuracy curve
exact_vals=[df2[f"top{k}_exact"].mean() for k in k_values]
concept_vals=[df2[f"top{k}_concept"].mean() for k in k_values]

plt.figure()
plt.plot(k_values, exact_vals, marker="o", label="Exact match")
plt.plot(k_values, concept_vals, marker="o", label="Concept match")
plt.xticks(k_values)
plt.xlabel("k")
plt.ylabel("Accuracy")
plt.title("Top-k Accuracy Curve (Case Level)")
plt.ylim(0,1)
plt.grid(True, linewidth=0.5, alpha=0.4)
plt.legend()
plt.savefig("topk_accuracy_curve.jpg", dpi=300, bbox_inches="tight")
plt.savefig("topk_accuracy_curve.pdf", bbox_inches="tight")
plt.show()


# %% [code]
# Category confusion matrix and category metrics (Top 1)
df2["gt_cat"]=df2["gt_primary"].apply(concept_key)
df2["pred_cat"]=df2["top1_pred"].apply(concept_key)

cats = sorted(set(df2["gt_cat"]) | set(df2["pred_cat"]))
cm = confusion_matrix(df2["gt_cat"], df2["pred_cat"], labels=cats)

plt.figure(figsize=(9,7))
plt.imshow(cm, aspect="auto")
plt.xticks(range(len(cats)), cats, rotation=45, ha="right")
plt.yticks(range(len(cats)), cats)
plt.title("Category Level Confusion Matrix (Top 1)")
plt.xlabel("Predicted category")
plt.ylabel("True category")
for i in range(cm.shape[0]):
    for j in range(cm.shape[1]):
        plt.text(j, i, str(cm[i,j]), ha="center", va="center", fontsize=8)
plt.colorbar()
plt.tight_layout()
plt.savefig("category_confusion_matrix.jpg", dpi=300, bbox_inches="tight")
plt.savefig("category_confusion_matrix.pdf", bbox_inches="tight")
plt.show()

report = classification_report(df2["gt_cat"], df2["pred_cat"], labels=cats, output_dict=True, zero_division=0)
cat_metrics = pd.DataFrame(report).T
cat_metrics


# %% [code]
# Most common category confusions
conf_pairs = (df2[df2["gt_cat"]!=df2["pred_cat"]]
              .groupby(["gt_cat","pred_cat"])
              .size()
              .reset_index(name="count")
              .sort_values("count", ascending=False))
conf_pairs.head(15)


# %% [code]
# Save a clean Excel summary
summary_overall = summary.copy()
by_img = df2.groupby("img_group").agg(
    n_cases=("echo_id","count"),
    top1_exact=("top1_exact","mean"),
    top5_exact=("top5_exact","mean"),
    top10_exact=("top10_exact","mean"),
    top1_concept=("top1_concept","mean"),
    top5_concept=("top5_concept","mean"),
    top10_concept=("top10_concept","mean"),
    mrr_concept=("mrr_concept","mean"),
    manual_contains=("manual_contains","mean")
).reset_index()

for c in ["top1_exact","top5_exact","top10_exact","top1_concept","top5_concept","top10_concept","manual_contains"]:
    by_img[c]=(by_img[c]*100).round(2)
by_img["mrr_concept"]=by_img["mrr_concept"].round(4)

with pd.ExcelWriter("MedGemma_Evaluation_Summary.xlsx", engine="openpyxl") as writer:
    summary_overall.to_excel(writer, index=False, sheet_name="Overall Metrics")
    by_img.to_excel(writer, index=False, sheet_name="By Image Count")
    pd.DataFrame(cm, index=cats, columns=cats).to_excel(writer, sheet_name="Category Confusion Matrix")
    cat_metrics.to_excel(writer, sheet_name="Category Metrics")
    conf_pairs.head(50).to_excel(writer, index=False, sheet_name="Common Confusions")
    df2.to_excel(writer, index=False, sheet_name="Case Level Results")

print("Saved: MedGemma_Evaluation_Summary.xlsx, topk_accuracy_curve.jpg/pdf, category_confusion_matrix.jpg/pdf")
```
