## Supplementary figures and images for "AI Decision Support for Challenging Teledermatology Cases: MedGemma Performance in the Dermatology ECHO Program"

### category_confusion_matrix_highres.png

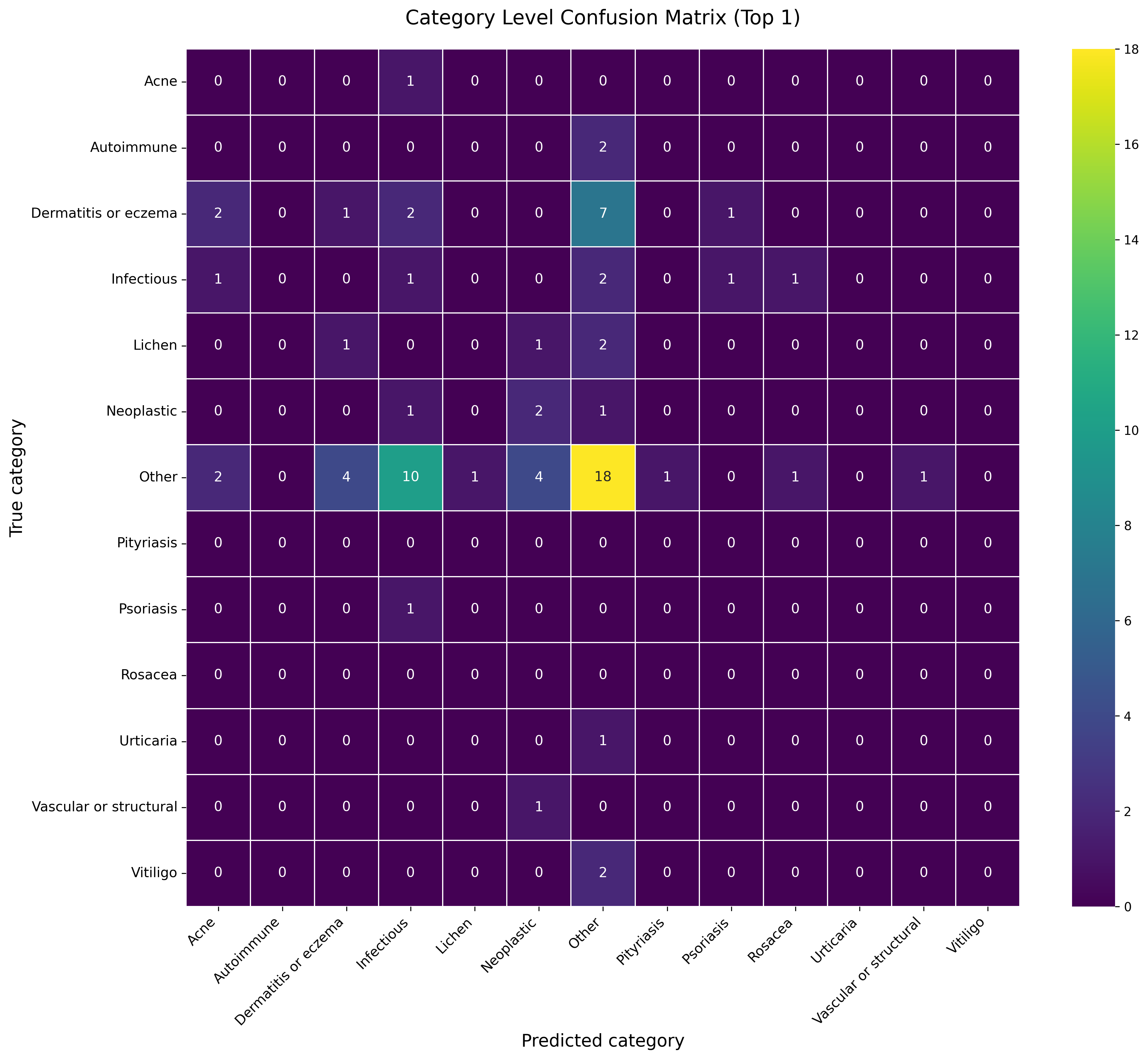

### full methods.png

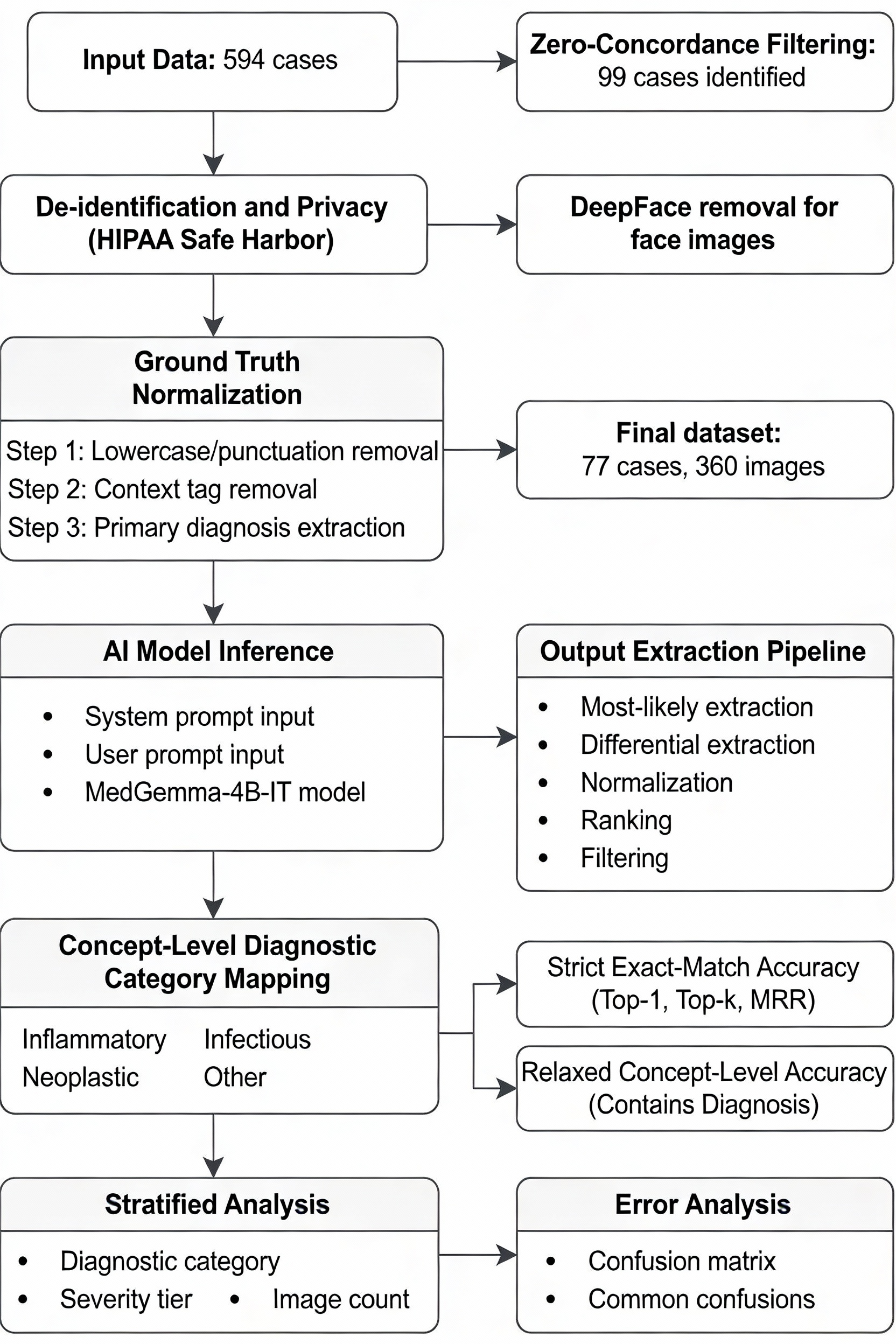

### topk_accuracy_curve.jpg

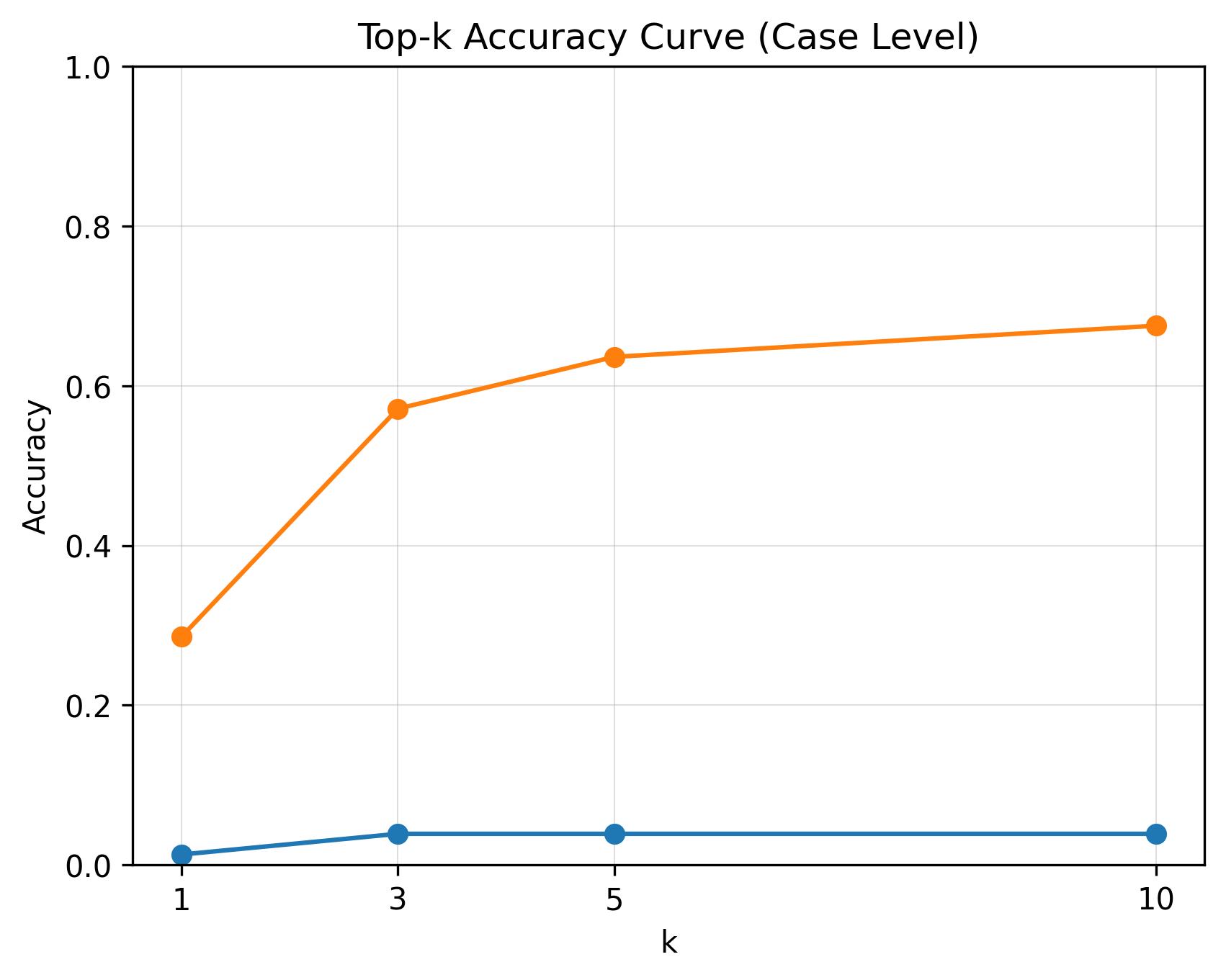
